# Linear plasmid prevalence and linezolid resistance gene carriage in vancomycin-resistant *Enterococcus* in Canada from 2009-2024

**DOI:** 10.64898/2026.05.08.26352429

**Authors:** Nicole Lerminiaux, Melissa McCracken, Jessica Bartoszko, Gurman Grewal, Sean Ahmed, Jennie Johnstone, George R Golding, the CNISP VRE working group

## Abstract

The incidence of vancomycin-resistant *Enterococcus* (VRE) is rising in hospitals in Canada, and resistance to last-resort antimicrobials including linezolid complicates treatment options for multidrug-resistant isolates. Recent reports from around the globe indicate that both linezolid and vancomycin resistance genes can be co-carried and mobilized by linear plasmids (named pELF) in *Enterococcus* species, often on the same backbone. We aimed to investigate linezolid resistance and linear plasmid prevalence in VRE bloodstream infection isolates collected by the Canadian Nosocomial Infection Surveillance Program from 2009 to 2024. We found that screening for pELF linear plasmid ends in short reads was a reliable way to predict linear plasmid presence in large-scale surveillance data (100 % accuracy on 85 reference samples). Almost half of the isolates in our collection were predicted to carry pELF plasmids (45.4 %, 941/2071) and we found that this proportion has increased from 2018 (32.2 %, 59/183) to 72 % of isolates between 2021 and 2024 (2021: 68.5 % (115/168); 2022: 71.6 % (146/204); 2023: 72.8 % (166/228); 2024: 71.6 % (235/328)). This trend of increasing linear plasmid carriage is evident from 2018 to 2024 across the dominant emerging sequence types (ST80, ST17, ST117). Linezolid resistance based on phenotypic antimicrobial susceptibility testing was low (1.0 %, 21/2071). Using long read sequencing, we characterized the linezolid resistant isolates and confirmed pELF plasmid presence in 13/21 (61.9 %) isolates. Six isolates harboured pELF plasmids encoding linezolid resistance genes (*optrA*, *cfr*(D), *poxtA*) and five of these also encoded vancomycin resistance genes (*vanA*). We compared these six plasmids to 39 public plasmid sequences and clustered them using MOB-suite and pling. Overall, this study provides further examples of the co-carriage of vancomycin and linezolid resistance genes on mobile linear plasmids and shows that linear plasmid prevalence is detectable and increasing across VRE in Canada.

**IMPACT STATEMENT:** Given the increasing prevalence of multidrug-resistant hospital-acquired pathogens, resistance to last-resort antibiotics is a global public health threat. Linezolid is a last-resort antibiotic used to treat vancomycin-resistant *Enterococcus* isolates, and the dissemination of linezolid resistance genes is significantly facilitated by mobile elements that can transfer between unrelated strains and species. Linezolid resistance genes have recently been described on linear plasmids and are often co-localized with other resistance genes on the same plasmid backbone. Consequently, understanding the features and distribution of linear plasmids and those harbouring linezolid resistance genes is crucial for pathogen surveillance and mitigation of resistance. In this work, we used long-read and short-read sequencing to characterize genomic epidemiology of linear plasmids across 16 years of *Enterococcus* surveillance data in Canada. This study furthers knowledge of linear plasmids by demonstrating that they are relatively common across vancomycin-resistant *Enterococcus* blood isolates and by providing more examples of co-localized vancomycin and linezolid resistance genes on the same linear plasmid backbone.

**DATA SUMMARY:** Sequencing data and genome sequences were deposited in National Centre for Biotechnology BioProject PRJNA1279082, and accessions are listed in **Table S1**. Supplementary materials for this study are available at the Figshare portal through DOI: XXX.

## INTRODUCTION

Vancomycin-resistant *Enterococcus* (VRE) are opportunistic pathogens associated with multidrug resistant hospital-acquired infections (1), with *Enterococcus faecium* and *Enterococcus faecalis* being the most clinically relevant species (2). Vancomycin-resistant *E. faecium* was listed as high priority on the World Health Organization’s 2024 bacterial priority pathogen list, indicating that they cause a substantial disease burden, are difficult to treat, and have increasing antimicrobial resistance (AMR) rates (3). According to the Canadian Nosocomial Infection Surveillance Program (CNISP), VRE bloodstream infection rates in Canada significantly increased from 0.30 to 0.37 per 10,000 patient days from 2019 to 2023 (4). *E. faecium* carrying the vancomycin resistance gene *vanA* is the most common type of VRE in Canada; *E. faecalis* and *vanB*-carrying isolates are rare (4).

Linezolid is a last-resort antimicrobial used to treat multidrug resistant VRE infections (2). Resistance to linezolid is emerging in VRE, which further limits treatment options for multidrug resistant infections (5). VRE can become resistant to linezolid via mutations in the 23S rRNA gene (G2576T or G2505A), mutations in ribosomal proteins L3 and L4, or by acquisition of AMR genes *optrA*, *poxtA*, and *cfr* variants (2,5). Linezolid resistance rates among *Enterococcus* were 1.6 % from 1997 to 2016 in the global SENTRY surveillance program (6) and varied from 1 to 5 % among national surveillance programs measured between 2013 and 2023 (7–13). Linezolid resistance among VRE bloodstream infections in Canada have been low (< 3 %) from 2019 to 2023 (4) and the genetic context of resistance has not been explored.

Both vancomycin and linezolid resistance genes are associated with mobile plasmids and transposons. The *vanA* genes are typically present on mobile plasmids that can transfer horizontally (14–17) and they are often associated with Tn*1546* and related structural variants (2,14). Acquired linezolid resistance genes are also found on conjugative plasmids in VRE and are associated with different transposons (18–22). For example, *optrA* is associated with Tn*6674* (22) and *poxtA* is associated with Tn*8082* (19).

Recently, linear plasmids (named pELF) have been described in *Enterococcus* species around the globe (7,12,19,20,23–33). These linear plasmids are typically between 70 to 160 kb in length and contain a hairpin structure at one end and a blunt invertron at the opposing end (24,33). pELF plasmids have a unique replicon gene (rep_pELF_) (30,33) and can transfer horizontally between *Enterococcus* species with self-encoded conjugation machinery (20,27,34). Furthermore, these linear plasmids have been implicated in multi-species nosocomial outbreaks (19,23,25,32). Vancomycin and linezolid resistance genes can be co-localized on the same pELF backbone (12,19,20,28,29,33), and this association has the potential to rapidly and simultaneously disseminate both types of AMR genes.

We aimed to characterize linezolid resistance and linear plasmid prevalence in the CNISP collection of VRE bloodstream infection isolates from 2009 to 2024. Using short-read (Illumina) sequencing, we investigated linear plasmid prevalence over time and how it relates to epidemiological data. Using long-read (Oxford Nanopore Technologies, ONT) sequencing, we examined the genomic context of linezolid resistance genes on linear plasmids in Canadian isolates and compared them to the global collection of enterococcal linear plasmids.

## METHODS

### Surveillance program and bacterial isolates

CNISP is a sentinel surveillance system that collects epidemiological and linked microbiology data from 109 Canadian acute care hospitals across 10 provinces and 1 territory (https://health-infobase.canada.ca/cnisp/). Eligible vancomycin-resistant isolates meeting surveillance case definitions (35) were submitted to the National Microbiology Laboratory (Winnipeg, Canada) for antimicrobial susceptibility testing and genomic characterization. From 2009 to 2024, 53 hospitals submitted 2071 *Enterococcus* bloodstream infection isolates from pediatric and adult inpatients with a minimum inhibitory concentration (MIC) to vancomycin of ≥8 μg/mL. Patients with a positive VRE blood isolate ≤14 days after completion of therapy for a previous VRE bloodstream infection, whether identified at the time of hospital admission or during hospitalization, were ineligible, along with emergency room and outpatient clinic cases not admitted to hospital. A patient could be included more than once in this study if a positive VRE blood isolate was identified >14 days after completion of therapy for a previous VRE bloodstream infection and was believed to be unrelated to the previous VRE bloodstream infection in accordance with best clinical judgement.

### Epidemiological data and statistics

Hospital staff trained in CNISP VRE bloodstream infection case criteria identified eligible patients, collected demographic and clinical data, including 30-day outcomes, using expert-reviewed and standardized questionnaires. Rates of VRE BSIs were calculated by dividing the total number of cases by the total number of patient days (multiplied by 10,000). We reported total numbers, frequencies, proportions and percentages for categorical variables and medians and interquartile ranges for continuous variables that were not normally distributed. Differences in proportions were tested using the χ2 or Fisher’s exact tests, while differences in medians were tested using the Wilcoxon rank sum test. All analyses were conducted in R version 4.5.0 at an alpha of 0.05.

### Antimicrobial susceptibility testing (AST)

Antimicrobial susceptibility testing was performed by broth microdilution using the GPALL1F Gram-positive Sensititre panels (Thermo Fisher Scientific, USA). Quality control strains ATCC29213 and ATCC29212 were tested concurrently in accordance with CLSI guidelines (36). MICs were interpreted according to CLSI breakpoints for all antimicrobial agents tested (36), with the exception of tigecycline which was interpreted according to EUCAST breakpoints (37).

### Whole-genome sequencing

All 2071 isolates were sequenced with Illumina platforms and isolates that were linezolid resistant (n=21) were additionally sequenced using ONT based on phenotypic resistance to linezolid identified by AST.

Genomic DNA was extracted using the DNeasy 96 Blood and Tissue kit (QIAGEN, Germany) for Illumina from 2009 to 2021 and the Mag-Bind® University Pathogen DNA 96 kit (Omega Bio-Tek, USA) for Illumina and ONT from 2022 to 2024. Short-read libraries were created with TruSeq NanoHT sample preparation kits (Illumina, USA) from 2009 to 2021 or the NexteraXT library preparation kits (Illumina) from 2022 to 2024. Paired-end, 301 bp indexed reads were generated on an Illumina MiSeq™ platform (Illumina) from 2010 to 2021 and a NextSeq 2000 platform (Illumina) from 2022 to 2024. Long-read sequences were generated using R10.4.1 chemistry on the Rapid Barcoding Kit (SQK-RBK114-24) (19/21) or R9.4.1 chemistry on the Rapid Barcoding Kit 96 (SQK-RBK110.96) (2/21) (ONT, UK). Read data were basecalled and demultiplexed with Dorado using the dna_r10.4.1_e8.2_400bps_sup@v5.2.0 model for R10.4.1 data or with Guppy v6.5.7 with the Super High Accuracy model (ONT) for R9.4.1 data. Average Illumina depth of coverage was 127 X, and average ONT depth of coverage was 346 X.

### pELF plasmid prediction using short reads and validation

We screened for pELF hairpin and invertron sequences in paired-end short reads using Bowtie2 v2.5.4 (38) to map reads to reference sequences and samtools v1.23 (39) to output the number of reads that mapped. Briefly, we constructed an index for the hairpin (position 0 to 799 of pELF1 sequence LC495616.1 (24) and invertron (position 142792 to 143316 of LC495616.1). Then we ran Bowtie2 with the ‘--very-sensitive-local --no-unal’ parameters, followed by samtools idxstats to output the number of reads that mapped per sample. More than five reads mapping at both the hairpin and invertron ends was considered a positive hit for pELF. We used a reference collection of VRE blood isolates from the National Microbiology Laboratory that are not part of CNISP for validation of this approach.

### Read processing and genome assembly

ONT reads were trimmed with Porechop v0.2.3_seqan2.1.1 (40) using default parameters and filtered for Q-score >10 and length >1,000 bases with nanoq (41) (parameters: -l 1000 -q 10). Illumina reads had adaptors trimmed and were filtered for an average Q-score >30 with trim-galore v0.6.7 (42) (parameters: --paired --quality 30). FastQC v0.11.9 (43) and NanoPlot v1.28.2 (44) were used to assess quality control metrics for Illumina and ONT reads, respectively. If average read depth for either Illumina or ONT reads exceeded 100X coverage, assuming a 3 Mb genome size, reads were randomly downsampled to 100X coverage with rasusa v2.0.0 (45). Isolates with Illumina-only data were assembled with Shovill v1.4.2 (46) (parameters: --depth 100 --nocorr --minlen 300 --mincov 2). Those with Illumina and ONT data were assembled with Autocycler v0.5.2 (47); if Autocycler assembly could not to resolve chromosomes (6/21), Hybracter v0.12.0 (48) (parameters: --skip_qc --medakaModel bacteria) was used. In addition, Flye v2.9.2 (49) (parameters: --nano-hq) was run on the Hybracter isolates for manual pELF hairpin end correction prior to polishing. Linear plasmids in Autocycler assemblies were cleaned using Bandage v0.8.1 (50) following the description in the Autocycler Wiki page and as described previously (51). All assemblies were rotated with Dnaapler v1.3.0 (52) and polished with Medaka v2.1.1 (53), Polypolish v0.5.0 (54), and Pypolca v0.3.1 (55).

### Bioinformatic analyses

StarAMR v0.11.0 (56) was used to detect antimicrobial resistance genes (ARGs) using the ResFinder database v2022-05-24 (57) and ST using the MLST database v2.23.0 (58). LRE-finder v1.0 was used to identify linezolid resistance mechanisms and copy number of mutations (59). Single nucleotide variants (SNV) were determined using SNVPhyl v1.2.3 (60) with the following parameters: minimum base coverage 10, minimum mapping quality score 30, relative SNV abundance ratio 0.75 and window size for SNV density filtering 20. SeqSphere+ software v9.0.10 (Ridom, Germany) was used to assess genetic relatedness using the *E. faecium* cgMLST scheme comprised of 1423 alleles (61). Bacteriocins were screened via blastn v2.14.0 (62) against a previously published database (63).

The MOB-typer tool from MOB-suite v3.1.9 (64,65) was used to identify plasmid replicons and mobility class using the default cutoffs and databases. Plasmid taxonomic unit (PTU) designations were obtained from COPLA v1.0 (66). Panaroo v1.3.2 (67) was used to estimate the pangenome and generate core gene alignments for the secondary clusters of interest. Pling v2.0 was used to determine the Double Cut and Join Indel (DCJ-Indel) distances between plasmids (68) and we tested DCJ Indel thresholds of 4 to 10 inclusive while keeping containment at the default 0.5. ISFinder (69) was used to identify inverted repeats for IS*1216E* and transposition activity was inferred by detecting 8-bp direct target repeats at the element’s boundaries.

For global plasmid clustering analysis, the PLSDB 2024_05_31_v2 database (70) was downloaded and clustered using MOB-suite as described above. Results were filtered for those in primary cluster AB369 and were clustered with pling as described above. In addition, we included linear sequences from recent pELF publications (12,19,26,31,71,72) and assembled four linear plasmids from (12) using the Hybracter/Flye hybrid assembly methods above.

## RESULTS

### Characteristics of VRE blood isolates

A total of 2071 non-duplicate VRE were submitted by 53 acute care hospitals across Canada from 2009 to 2024. Most isolates encoded *vanA* (94.6 %, 1959/2071), followed by *vanB* (5.2 %, 106/2071), or both *vanA* and *vanB* (0.3 %, 6/2071). None of these isolates encoded daptomycin resistance operon *drcAB*.

Only 16 isolates were *Enterococcus faecalis*, all of which encoded *vanA*, and the remaining isolates were identified as *Enterococcus faecium*. Altogether, these isolates comprised 91 distinct STs and 505 unique cgMLST types (**Supplementary Figure 1**), with the most common STs being ST117 (21.7 %, 449/2071), ST80 (18.2 %, 377/2071), ST117 (17.5 %, 362/2071), and ST1478 (11.1 %, 230/2071). The dominant sequence types from 2018 to 2024 are illustrated in **Fig. 1A**, with the most common sequence types over this period being ST1478, ST17, ST117, and ST80. In 2018, the dominant ST in Canada was ST1478; this type began to decline in 2019 and was replaced by ST17 and ST80. Since 2023, ST117 has increased in proportion and may become the next dominant ST.

**Figure 1.**
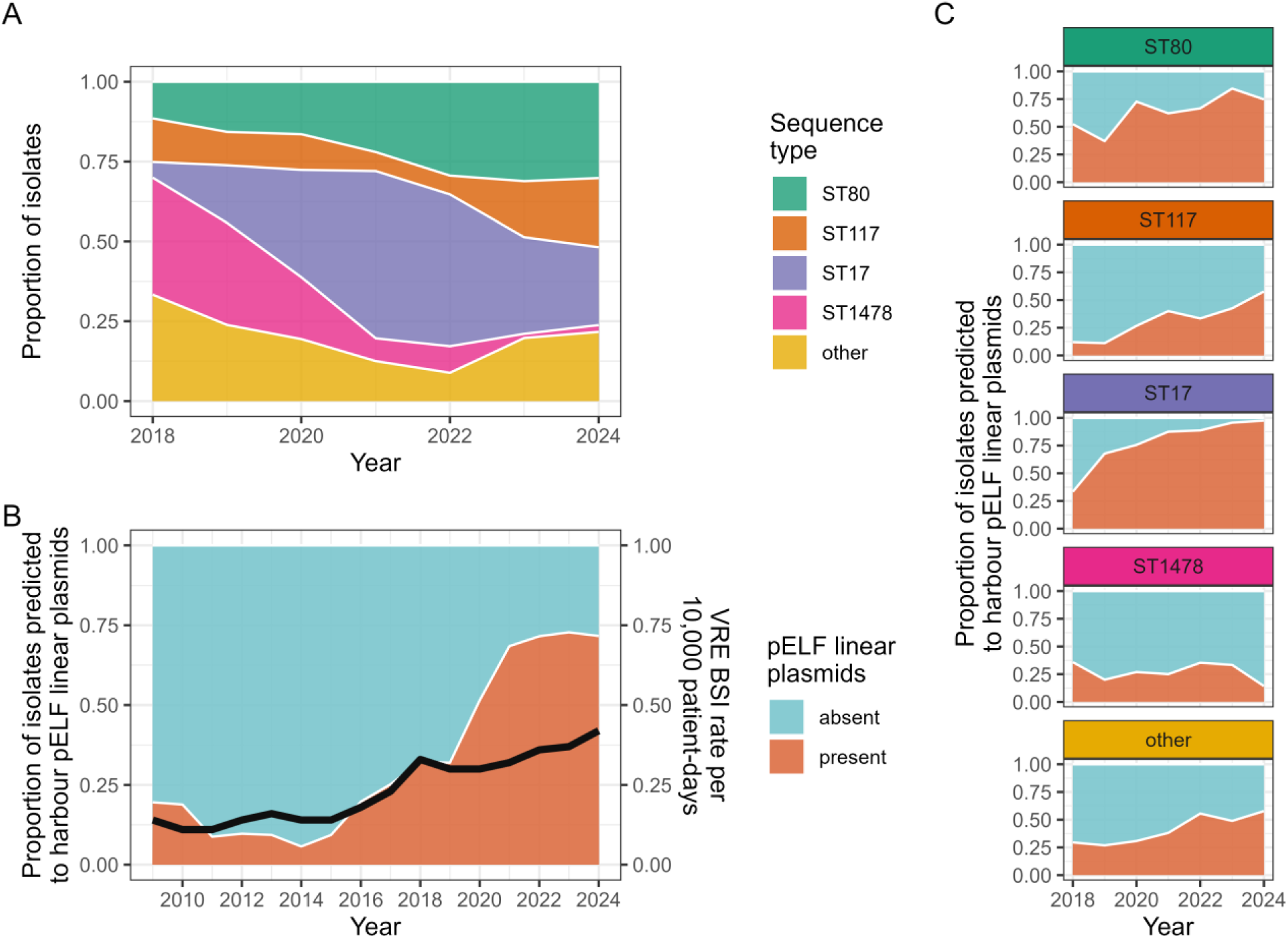
(**A**) Dominant VRE sequence types from 2018 to 2024 over time. This plot represents all isolates in the collection, including those with and without predicted pELF linear plasmids. (**B**) Proportion of total VRE isolates from 2009 to 2024 predicted to harbour pELF linear plasmids overlaid with the VRE bloodstream infection rate per 10,000 patient days (black line). (**C**) Proportion of isolates from dominant VRE sequence types from 2018 to 2024 harbouring a predicted pELF linear plasmid.

### pELF plasmid prevalence is increasing over time and mirrors the rise and fall of distinct STs

We performed short-read sequencing on all isolates and screened our collection for the linear plasmid pELF1 hairpin and invertron sequences (accession: LC495616.1) as others have done previously (7). We validated this approach using the 21 isolates sequenced with ONT here and an additional 64 isolates from a reference collection (n=85, see Methods for details) and found it was 100 % accurate; those predicted to have pELF linear plasmids using this approach (n=51) did indeed have pELF linear plasmids confirmed by sequencing and assembling ONT data, and those that were not predicted to have pELF linear plasmids (n=34) did not have a linear plasmid assembled with ONT data (data not shown). We attempted aligning assembled contigs from short-read Shovill assemblies to the reference hairpin and invertron sequences to predict linear plasmid presence on a subset of this validation dataset (n=24), however those sequences were often missing from assembled contigs and pELF linear plasmid presence was correctly detected in only 50 % of isolates. In addition, we assessed whether MOB-recon could accurately predict pELF plasmids by reconstructing short-read contigs into the main pELF secondary clusters identified in this study (AK115, AK116, AK117) using the same subset and this approach was only 80% accurate.

Almost half of the isolates in our collection were predicted to harbour a pELF linear plasmid (45.4 %, 941/2071), and we found the prevalence of pELF plasmids increased in VRE over time (**Fig. 1B**). The CNISP VRE collection dates back to 1999 but is not comprehensive prior to 2009; nevertheless, the first detected linear plasmid in our collection was in 2002. In 2009, the proportion of VRE isolates with a pELF plasmid was 19.6 %; this dropped to around 5 to 10 % from 2010 to 2015, and then began increasing up to 2021, where the rate has since stabilized around 72 %. The increase in VRE bloodstream infection rate per 10,000 patient days has steadily increased from 0.14 in 2009 to 0.42 in 2024.

We examined pELF prevalence in the dominant sequence types (STs) and found that the proportion of VRE carrying pELF increased across most since 2018 (**Fig. 1C**). Prevalence was very high for ST17 in 2024 (97.4 %, 76/78) and increased in ST117 from 12.0 % (3/22) in 2018 to 58.8 % (40/68) in 2024. In contrast, the proportion of pELF prevalence in ST1478 has decreased from 35.8 % (24/67) in 2018 to 14.3 % (1/7) in 2024.

We analyzed clinical epidemiological data to determine if there were differences between VRE blood isolates predicted to carry pELF compared to those without using data from 2018 onwards (**Table 1**). We found that isolates from the West region were significantly more likely to harbour linear plasmids, while isolates from the Central region were significantly less likely (p < 0.001). A history of solid organ transplant was significantly less common among VRE bloodstream infection patients with linear plasmids compared to VRE BSI without linear plasmids (p < 0.001). Linezolid non-susceptibility was slightly more common among isolates with linear plasmids compared to those without, but the difference was not statistically significant.

**Table 1.** Patient characteristics and clinical outcomes by VRE bloodstream infection isolate linear plasmid status from 2018 to 2024.

| Characteristic | Linear plasmid present<br>N=821<sup>1</sup> | Linear plasmid absent<br>N=557<sup>1</sup> | p-value |
| --- | --- | --- | --- |
| **Region<sup>2</sup>** |  |  | **<0.0013** |
| Central | 336/821 (41%) | 285/557 (51%) |  |
| East | 5/821 (0.6%) | 1/557 (0.2%) |  |
| West | 480/821 (58%) | 271/557 (49%) |  |
| **Age (years)** |  |  | 0.74<sup>4</sup> |
| Mean (SD) | 59 (17) | 58 (18) |  |
| Median (Q1, Q3) | 62 (50, 71) | 61 (50, 71) |  |
| Min, Max | 0, 97 | 0, 94 |  |
| **Biological sex** |  |  | 0.51<sup>5</sup> |
| Female | 315/821 (38%) | 204/557 (37%) |  |
| Male | 506/821 (62 %) | 353/557 (63%) |  |
| **Hemodialysis** | 275/743 (37%) | 186/494 (38 %) | 0.82<sup>5</sup> |
| **Chemotherapy** | 146/743 (20%) | 108/494 (22%) | 0.35<sup>5</sup> |
| **Bone marrow or stem cell transplant recipient** | 46/749 (6.1%) | 44/508 (8.7%) | 0.089<sup>5</sup> |
| **Solid organ transplant recipient** | 56/766 (7.3%) | 67/504 (13 %) | **<0.001<sup>5</sup>** |
| **Linezolid treatment** | 268/746 (36%) | 167/498 (34%) | 0.39<sup>5</sup> |
| **Daptomycin treatment** | 378/746 (51%) | 253/498 (51%) | 0.96<sup>5</sup> |
| **Resistance to linezolid** |  |  | 0.14<sup>5</sup> |
| Non-susceptible | 54/821 (6.6%) | 26/557 (4.7%) |  |
| Susceptible | 767/821 (93%) | 531/557 (95%) |  |
| **Resistance to daptomycin** |  |  | 0.82<sup>5</sup> |
| Non-susceptible | 29/821 (3.5%) | 21/557 (3.8%) |  |
| Susceptible | 792/821 (96%) | 536/557 (96%) |  |
| **30-day ICU admission** |  |  | 0.052<sup>5</sup> |
| Already in ICU | 241/791 (30%) | 145/515 (28%) |  |
| Yes | 104/791 (13%) | 93/515 (18%) |  |
| No | 446/791 (56%) | 277/515 (54%) |  |
| **30-day all-cause mortality** | 276/819 (34%) | 187/551 (34%) | 0.93<sup>5</sup> |

<sup>1</sup>n/N (%). Note variation in denominators reflects differences in missingness (i.e. unknown or incomplete responses)
<sup>2</sup>Central: Ontario, Quebec. East: New Brunswick, Nova Scotia, Newfoundland and Labrador, Prince Edward Island. West: British Columbia, Alberta, Saskatchewan, Manitoba.
<sup>3</sup> Fisher's exact test
<sup>4</sup> Wilcoxon rank sum test
<sup>5</sup>Pearson's Chi-squared test

### Linezolid resistance in Canada is low

Of the VRE screened in our collection, 21 *E. faecium* were resistant to linezolid (MIC ≥8 ug/mL) by phenotypic AST (1.0 %, 21/2071) (hereafter referred to as LVREfm) (**Table S1**). Using short-read assemblies, we found that the majority (71.4 %, 15/21) had G2576T mutations in the 23S rRNA genes (known to cause a resistant phenotype (73)), and the remainder encoded *cfr*(D)*, optrA*, and/or *poxtA* genes (28.6 %, 6/21) (**Fig. 2**). Out of 6 total 23S rRNA gene copies, the LVREfm with G2576T mutations encoded between 1 and 4 mutated 23S rRNA copies. A single mutated 23S rRNA gene is enough to produce a resistant phenotype (59), and we observed that more copies of the mutated 23S rRNA gene resulted in a higher linezolid MIC value (MIC = 8 µg/mL for 1-2 copies vs MIC > 8 µg/mL for 3-4 copies). Along with the VanHAX operon present in all LVREfm isolates, other common antimicrobial resistance genes included *msr*(C) (100 %, 21/21), *aac*(6’)-Ii (100 %, 21/21), *tet*(M) (95.2 %, 20/21), and *erm*(B) (81.0 %, 17/21). These isolates comprised 8 STs and 11 cgMLST types, with the most common STs being ST80 (28.6 %, 6/21), ST17 (23.8 %, 5/21), and ST1478 (19.0 %, 4/21). Several of these isolates appeared to be clonally related (05A21018 & 05A21021: 12 SNVs over 88 % core, 07D18011 & 07D18014 & 08A19002: < 5 SNVs over 88 % core).

**Figure 2.**
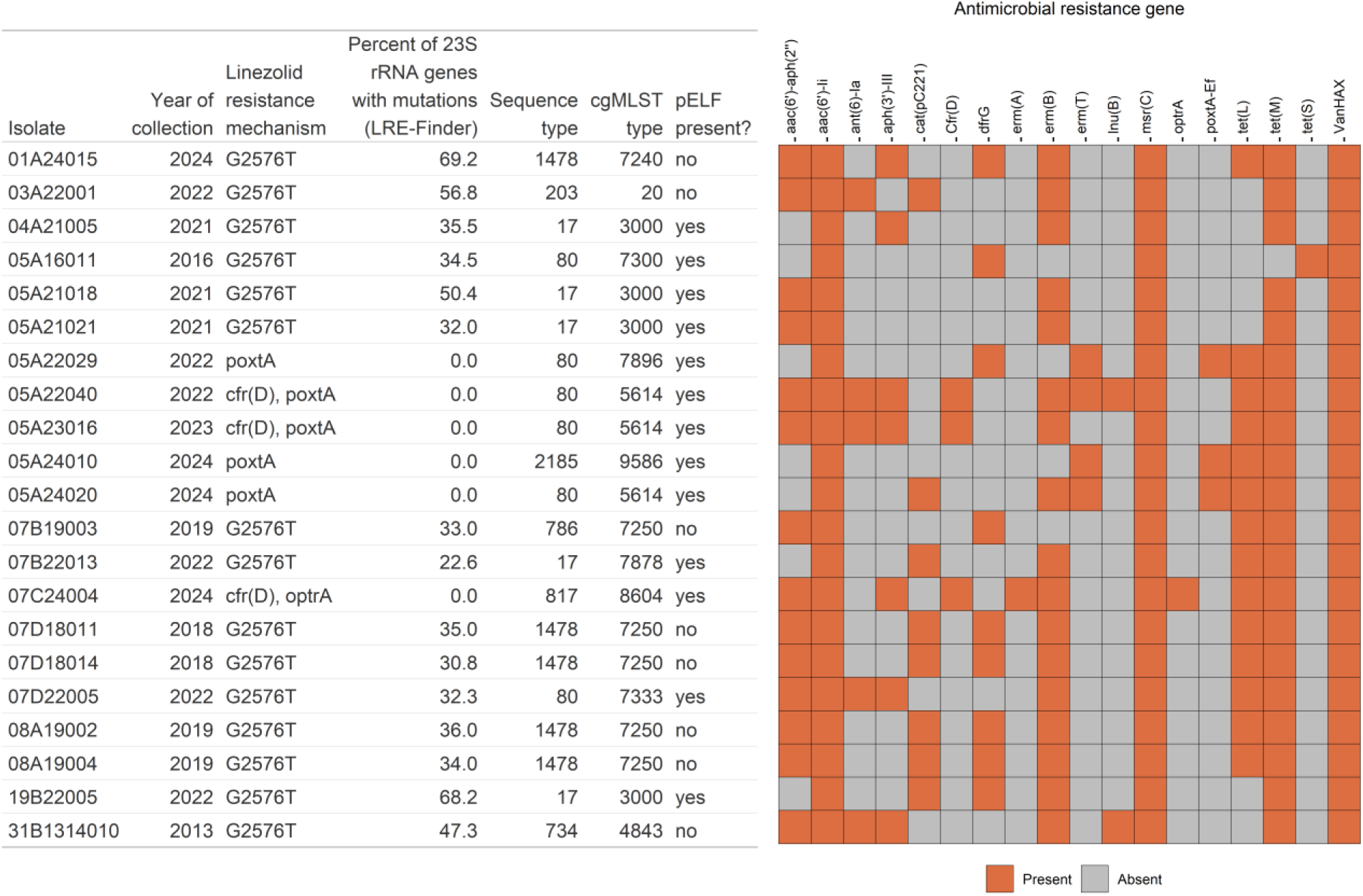
Metadata and AMR gene heatmap for 21 linezolid-resistant VRE isolates in the CNISP collection.

Epidemiologically, these isolates were found in both the West region (n=12) and Central region (n=9) and were found in both male (n=14) and female (n=7) patients. The average age was 55 years (range: 22 – 75). Phenotypic AST profiles were identical for ampicillin (resistant, MIC > 8 µg/mL), penicillin (resistant, MIC > 8 µg/mL) and vancomycin (resistant, MIC ≥ 32 µg/mL); full AST results can be found in **Table S2**.

### Linezolid resistance genes are carried on pELF plasmids in Canada

Given the recent reports of linezolid resistance genes on pELF plasmids globally (7,12,19,20,28,33) and in Canada (29), we sequenced all 21 LVREfm isolates with ONT to assess linear plasmid presence. Based on screening for the pELF hairpin and invertron sequences in short reads as described above, most of the isolates (61.9 %, 13/21) were predicted to contain linear pELF-type plasmids. This was confirmed with ONT assembly, wherein the same 13 isolates identified did have linear pELF-type plasmids ranging from 88 to 126 kb in size, with one exception at 330 kb (**Fig. 3**). In addition, the 8 LVREfm isolates which were not predicted to harbour a linear plasmid in short reads did not have a linear plasmid assembled, which indicates that short-read mapping to hairpin and invertron sequences is a reliable method for screening. Following the pELF nomenclature (7,24), we named these plasmids with a pELF prefix. With the exception of a large 330 kb plasmid (pELF_04A21005), all LVREfm pELF plasmids grouped in the same MOB-suite primary cluster (AB369) and in several secondary clusters, with the most common being AK115 (69.2 %, 10/13). COPLA classified all 13 LVREfm pELF plasmids as PTU-Lab16. A plasmid replicon gene *rep*_pELF_ (30,33) and a conjugative *tra* operon (34) have recently been described in pELF linear plasmids; all 13 LVREfm pELF plasmids encoded the *rep*_pELF_ gene at > 95 % nucleotide identity and the conjugative *tra* operon > 96.9 % nucleotide identity.

**Figure 3.**
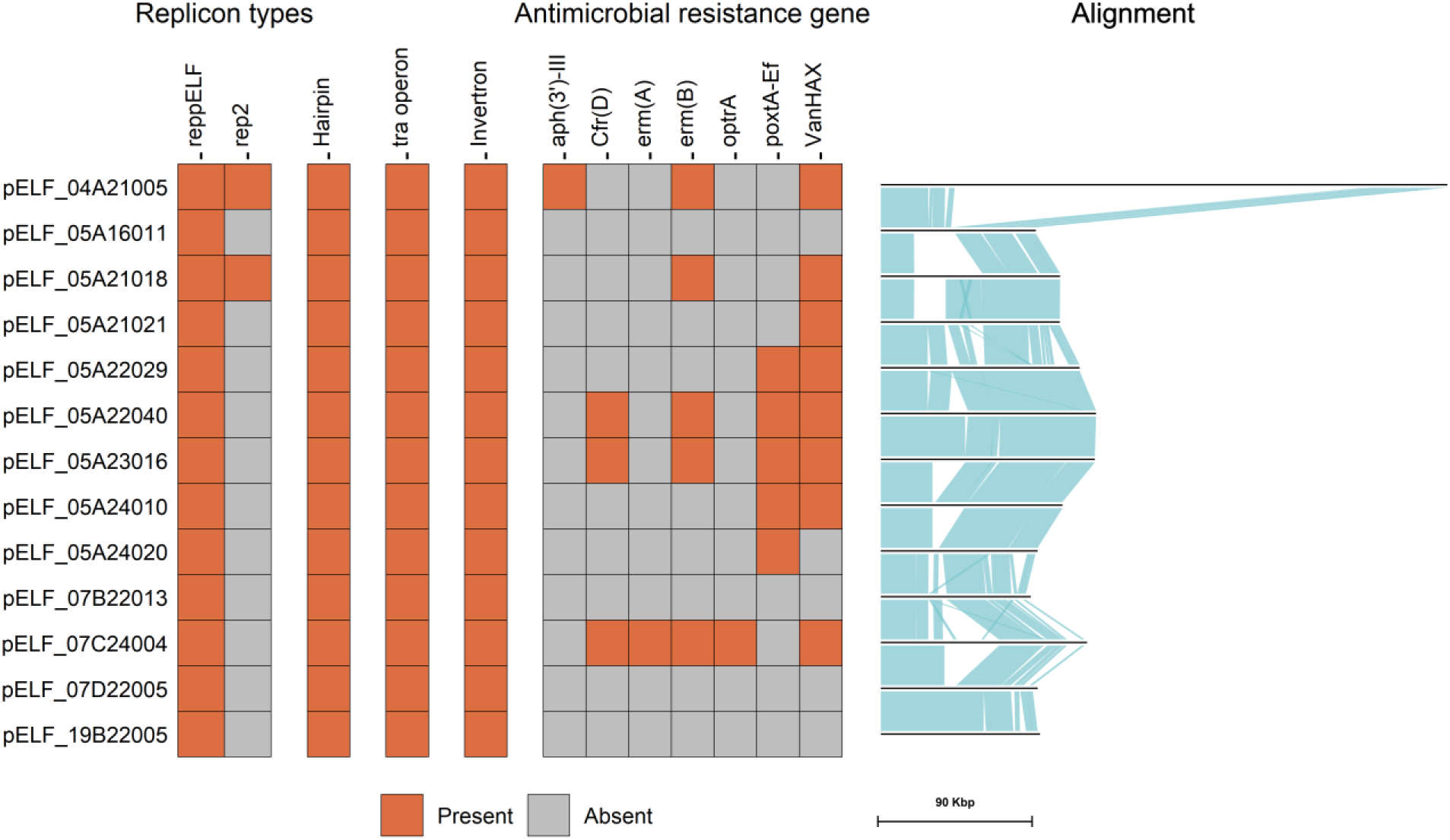
Heatmap of features found on pELF linear plasmids in linezolid and vancomycin-resistant Canadian isolates (n=13). Right panel: pairwise blast identity between sequences, with blue blocks representing homology > 85 % over 1000 bases.

One pELF plasmid was considerably larger than the others. pELF_04A21005 was 330.5 kb in length and contained approximately 237 kb (∼70 %) of sequence situated between the rep_pELF_ replicon and the *tra* conjugative operon that was unique compared to other pELF plasmids from Canada and those publicly available in NCBI (**Fig. 4**). The insertion was flanked on either side by IS*6* family IS*1216E* transposases with a AAAATGGA direct repeat, indicative of transposase-mediated insertion in the pELF backbone. This plasmid contained a total of nine copies of IS*1216E*. The inserted sequence had 84 % coverage and > 99 % nucleotide identity with a 222 kb circular plasmid from a clinical VRE isolate in NCBI (CP197034.1) but had undergone structural rearrangement. It encoded VanHAX, *erm*(B), *aph*(3’)-III, a rep2 replicon, a MOBP relaxase and a MPF_T mating pair formation protein, and it was predicted to be conjugative by MOB-suite.

**Figure 4.**
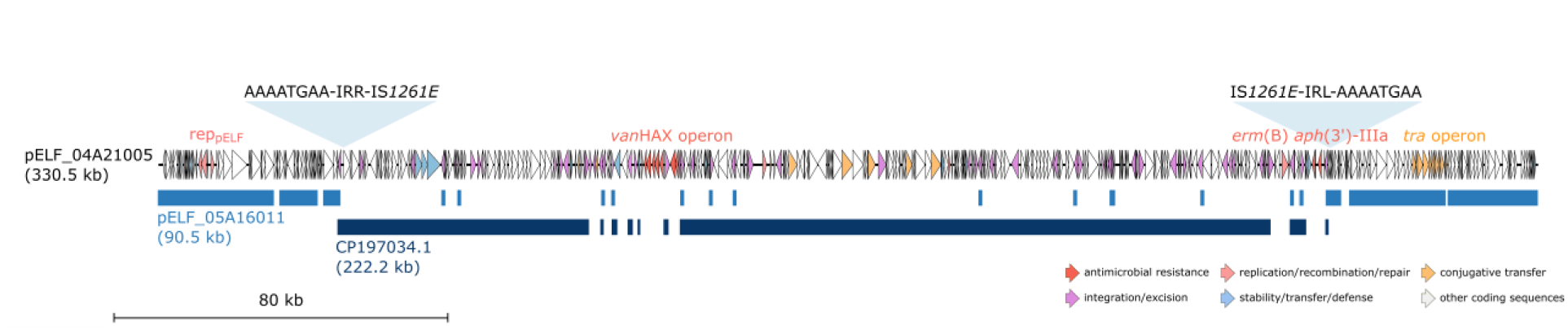
Map of linear plasmid pELF_04A21005 (330.5 kb). Genes are coloured by function, and not all labels are displayed. Similar plasmid sequences (CP197034.1: 222.2 kb and pELF_05A16011: 90.5 kb) were included to show integration into the pELF backbone. Transposon insertion sites mediated by IS*1261E* including the inverted repeats (right: IRR, left: IRL) and target site duplication sequences (AAAATGAA) are depicted above the backbone. The hairpin is on the left side and the invertron is on the right.

Six LVREfm isolates encoded *cfr*(D), *optrA* and/or *poxtA* on the pELF linear plasmids, with one isolate (05A24020) encoding a second copy of *poxtA* on the chromosome. Of these, 5/6 also encoded VanHAX on the same pELF plasmid. The other LVREfm isolates (n=7) with pELF plasmids that did not carry acquired linezolid resistance genes harboured either VanHAX and *erm*(B) on the pELF plasmids (23.1 %, 3/13) or no AMR genes at all (30.8 %, 4/13). No bacteriocins were detected on any of the pELF plasmids.

We examined the genetic context of the linezolid and vancomycin resistant genes on the pELF plasmids (**Supplementary Figure 2**). All 3 pELF plasmids with *cfr*(D) had this gene located between two IS*1261E* copies as described previously for pELF plasmids (20). The single pELF plasmid encoding *optrA* had a truncated version of *Tn6674* which includes *optrA* downstream of IS*256* which has been described previously (20). All 5 pELF plasmids encoding *poxtA* had 100% nucleotide identity or 1 SNV over the 8.2 kb Tn*8026* [Hall et al 2026]. The isolate with two *poxtA* copies (one chromosomal and one on pELF) had two identical copies of Tn*8026*. All 8 pELF plasmids encoding VanHAX had these genes located in a truncated Tn*1546* transposon as described previously for pELF plasmids (19,20,28).

### Comparing Canadian lzdR linear plasmids to global lzdR linear plasmids

We examined other pELF plasmids in NCBI and PLSDB that grouped in the same MOB-suite primary cluster AB369 or that appeared in recent pELF literature (n=154, **Table S3**). Several of the plasmid sequences appeared to be incomplete (missing the hairpin or invertron sequences, extra complementary sequence past the hairpin, or were fragmented in two pieces) or were annotated as circular in NCBI. In addition, we assembled 4 plasmids with public ONT data (12) using the same methods as above. Most plasmids grouped in MOB-suite secondary cluster AK117 (50.6 %, 78/154), followed by AK115 (31.2 %, 28/154) and AK116 (8.4 %, 13/154) (**Table S3**). Using a DCJ-Indel distance of 10, pling grouped most plasmids into 4 of 10 subcommunities (39.6 % 61/154; 32.5 % 50/154; 9.7 % 15/154; and 8.4 % 13/154) (**Table S3**). Over half encoded at least one AMR gene (68.1 %, 105/154), with 95 encoding a *van* gene cluster (61.7 %, 95/154) and 39 encoding linezolid resistance genes *cfr*(D), *optrA,* and/or *poxtA* (25.3 %, 39/154). Additional AMR genes frequently observed were *erm*(B) (38.3 %, 59/154), *aph*(3’)-III (14.9 %, 23/154), *erm*(A) (14.2 %, 22/154), and *ant*(9)-Ia (7.8 %, 12/154). The most common AMR genotype of pELF plasmids was VanHAX, *cfr*(D), erm(B), *poxtA* (15.6 %, 24/154), followed by VanHAX (14.3 %, 22/154), then VanHAX, *ant*(9)-Ia, and *erm*(A) (5.2 %, 8/154), and VanHAX, *aph*(3’)-III, and *erm*(B) (5.2 %, 8/154). Each of the main MOB-suite secondary clusters and pling subcommunities contained pELF plasmids with no AMR genes.

We compared the 39 LVREfm pELF plasmids from the global collection with the 6 from our Canadian collection (n=45, **Fig. 5**). All pELF plasmids were isolated from human sources, except one from sewage, and the collection dates ranged from 2017 to 2024. About half of these (n=24) were from a hospital-associated outbreak of ST80 in Australia (19). There were 8 STs represented, and most isolates were ST80 (75.6 %, 34/45). All plasmids encoded the rep_pELF_ gene and the conjugative *tra* operon. Most plasmids were grouped into secondary cluster AK115 by MOB-suite (93.3 %, 42/45). Three plasmids (6.7 %, 3/45) were hybrid co-integrates; they grouped in different secondary clusters, they encoded additional replicon types, and they were larger in size (> 130 kb). The linezolid resistance genes *cfr*(D), *optrA*, and *poxtA* were found on 80.0 % (36/45), 26.7 % (12/45), and 75.6 % (34/45) of plasmids, respectively. Other common AMR genes on these plasmids were VanHAX (88.9 %, 40/45), *erm*(B) (82.2 %, 37/45), and *erm*(A) (24.4 %, 11/45).

**Figure 5.**
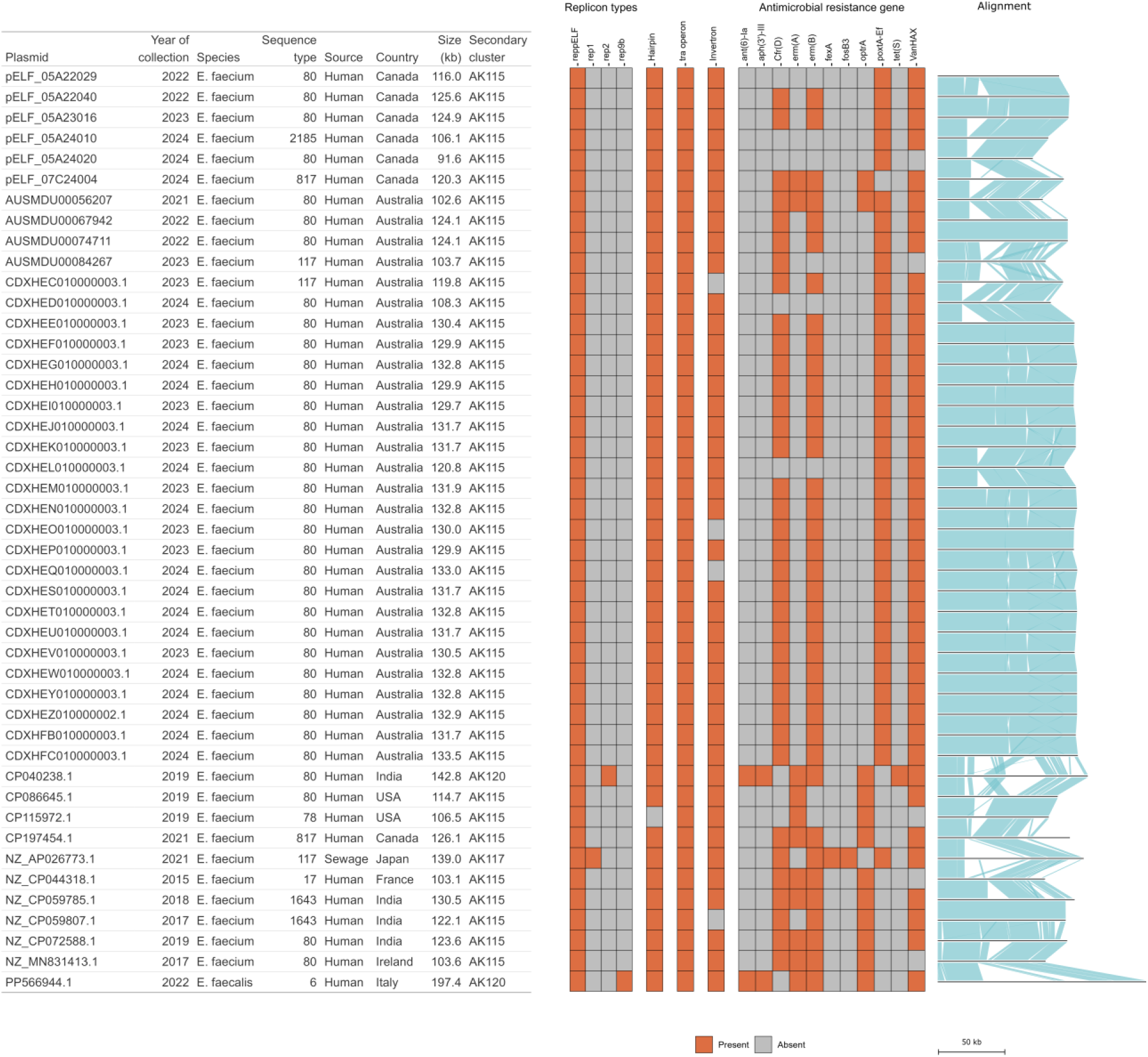
Metadata and heatmaps for pELF plasmids carrying linezolid resistance genes from this study (n=6) and others (n=39).

We examined plasmid relatedness of cluster AK115 (n=42) using pling and computed a pangenome, and excluded those plasmids classified into different MOB-suite secondary clusters (n=3) (**Fig. 6**). Of the remaining 42 LVREfm pELF plasmids, the pangenome comprised 215 genes, of which 29.7 % (64/215) were core genes (one copy present in every plasmid), and 46.5 % (100/215) were soft core genes present in at least 90 % of plasmids. These core genes included replicon proteins, the twelve genes in the conjugative *tra* operon, toxin-antitoxin systems, and hypothetical proteins (**Table S4**). Pling clustering identified 9 subcommunities: five singletons, two groups of two plasmids, one group of 10, and one group of 23. The mean number of structural changes (DCJ-Indel distances) between plasmids was 8.3, indicating structural diversity among this plasmid group. The largest subcommunity (labeled I in **Fig. 6**) contained sequences from a clonal outbreak in Australia (19). The second largest subcommunity (labeled H in **Fig. 6**) containing 10 plasmids were isolated from Canada (this study), India, and Australia. Six of these were from ST80 hosts but isolated several years apart, and SNV analysis indicated their host genomes were not closely related (average SNV distance: 41, data not shown). The two subcommunities containing two plasmids (labeled F and G in **Fig. 6**) were previously described as having similar sequences (7,29). Changing the default DCJ-Indel threshold from 4 to 5 resulted in the combination of subcommunities I, H, and E into one. Ampicillin

**Figure 6.**
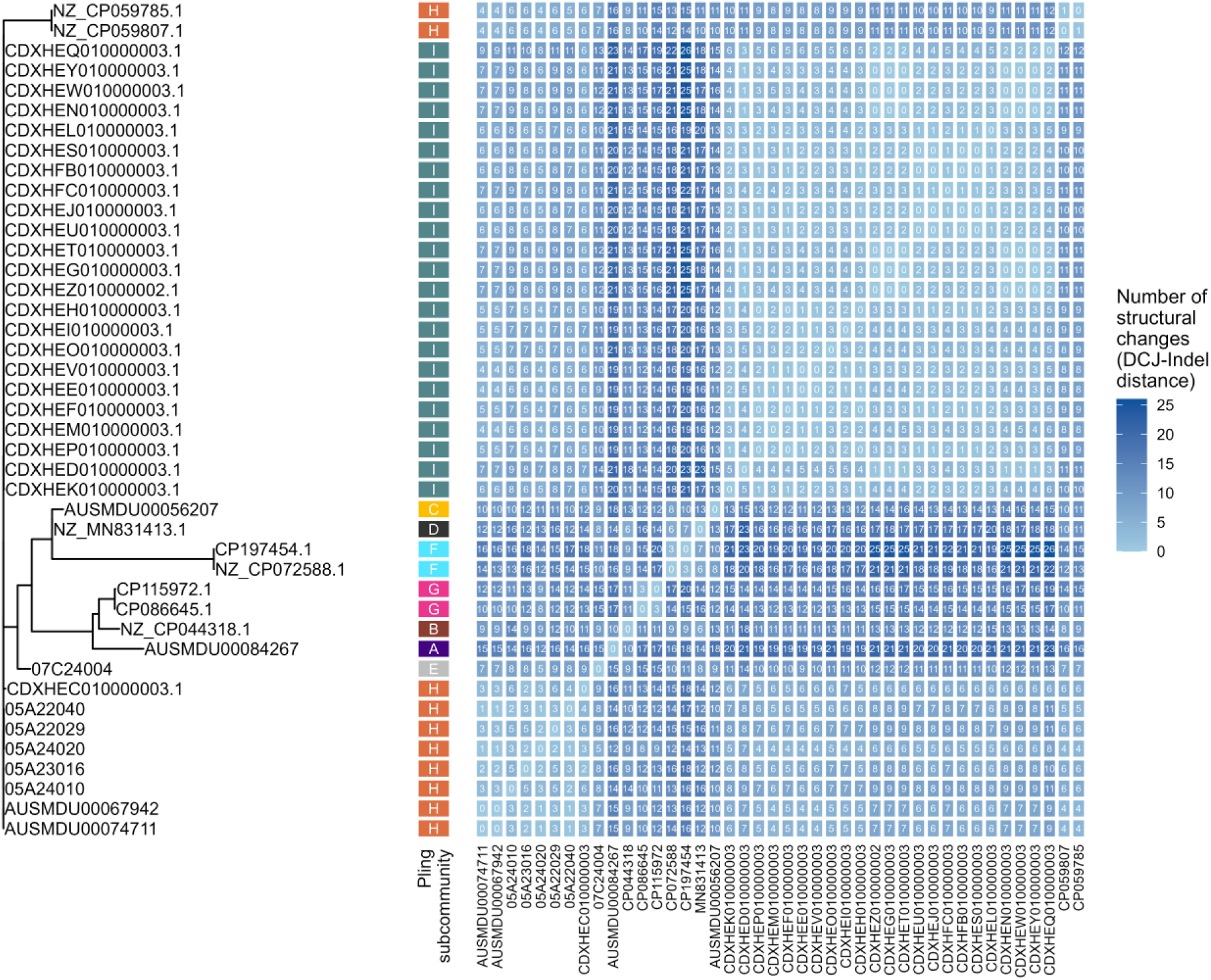
Core gene alignment of 42 pELF linear plasmids carrying linezolid resistance genes and heatmap of pling DCJ-Indel distances. Cores genes were defined as one copy present in each plasmid (64/215 total genes). DCJ-Indel distances represent the lowest number of structural changes between each plasmid pair. Pling defined subcommunities using a containment threshold of 0.5 and a DCJ-Indel threshold of 4.

## DISCUSSION

Linear plasmids (pELF) in VRE can encode multiple genes for resistance to last-resort antimicrobials vancomycin and linezolid. Recent reports indicate linear transferable mobile elements are involved in linezolid resistance dissemination (12,19,20,28,29,33). While much is still unknown about the biology of pELF linear plasmids, the co-localization of AMR genes on these mobile elements has potential for rapid horizontal dissemination across strains and species, making this a global public health threat. Here we described the prevalence of pELF plasmids across VRE blood isolates from a national surveillance program in Canada (2009–2024) and demonstrated that pELF prevalence has increased over time across dominant sequence types. We examined the linezolid resistant isolates (n=21) in this collection and characterized their linear plasmids. Six isolates (46.2 %) encoded linezolid resistance genes on pELF plasmids and 5/6 (83.3 %) also encoded vancomycin resistant genes co-localized on the same plasmid. We compared these plasmid sequences to the global collection of pELF plasmids and highlighted similarities through clustering.

The level of linezolid resistance in Canada was comparable to that reported by surveillance programs in other countries. The USA reported linezolid resistance in 2.0 % of *E. faecium* and 0.6 % of *E. faecalis* from 2018 to 2019 (7), which was slightly higher compared to the 1.0 % of VRE found here. The global SENTRY surveillance program observed 1.6 % of VRE from 1997 to 2016 were resistant to linezolid, and proportions were within 1 % across all continents (6). The EU reported linezolid resistance in 1.6 % of VRE from 2014 to 2018 (8), and this agreed with Switzerland (1.4 % of *E. faecium* from 2013 to 2018, (9)) and Denmark (1.1 % of VRE from 2015 to 2022 (10)). Several countries reported slightly higher levels of resistance; China found linezolid resistance in 3.5 % of *E. faecalis* and 0.6 % of *E. faecium* in 2022 (11), Australia had linezolid resistance in 4.3 % of *E. faecium* and 0.3 % of *E. faecalis* from 2015 to 2023 (12), and Egypt reported linezolid resistance in 5.2 % of VRE (13). The G2576T mutations in the 23S rRNA genes were the most common mechanism of linezolid resistance observed here and in other countries including the USA (7), Belgium (74), Denmark (10), Germany (75), and Japan (76). G2576T mutations were more common in *E. faecium* than *E. faecalis* in the SENTRY surveillance program (77), and *optrA* was more common in *E. faecalis* whereas *cfr*(D), *optrA* and *poxtA* were similarly common in *E. faecium* and we often co-carried in a global dataset (12). We did not detect any linezolid-resistant *E. faecalis* in our VRE collection. The incidence of linezolid resistance appears relatively low in Canada and resistance mechanisms agree with previously published results.

The overall proportion of VRE predicted to encode pELF-type plasmids observed in the Canadian collection is much higher than previous studies. Hashimoto *et al*. (25) identified 2.6 % of VRE had pELF-like plasmids in their Japanese collection from 2000 to 2019. Kent *et al*. (7) observed that 20 % of VRE were predicted to carry pELF plasmids in their *E. faecium* USA surveillance dataset collected from 2018 to 2019. In our Canadian dataset, the proportion of VRE with pELF plasmids jumped from 32 % in 2018 to 51 % in 2020 and continued to increase to around 72 %. This increase may also be found in other countries during the same time frame, but this has not yet been studied to the best of our knowledge. Among our collection from 2018 to 2024, linezolid resistance was not significantly more likely to occur in isolates with linear plasmids. This is something worth observing over future years to see if this trend does become statistically significant, as the incidence of bloodstream VRE infections that harbour linezolid resistance on linear plasmids has increased over time.

The increase in pELF prevalence in 2021 mirrors the rise of certain sequence types that had a high proportion of isolates with pELF. There have been ongoing shifts in dominant VRE clones globally and even at specific hospital sites (1); the downfall of ST1478 coincides with an increase in pELF prevalence across ST117 and ST80 around the globe (10,78–81). However, it is difficult to determine if pELF possession favoured host expansion relative to those without, or if pELF prevalence increased due to successful host expansion attributed to other factors. The timing of increasing pELF prevalence also coincides with an overall increase in VRE rates observed in multiple countries (reviewed in (1)). The increase in VRE from 2020 may be due to limited resources during COVID-19, including healthcare facilities stretched to the limits, longer hospital stays, difficulty implementing infection prevention and control practices, and misuse of antibiotics (82,83). Perhaps these factors during 2020 to 2021 favoured expansion of hosts with pELF plasmids.

One feature of pELF plasmids that may promote their expansion are addiction modules encoded on the plasmids. A recent study described an increase in plasmid-borne bacteriocin prevalence in *E. faecium* from 2016 to 2021 with a particular association in vancomycin-resistant isolates and dominant sequence types 117 and 80 (84). The authors discussed how this bacteriocin provided isolates with a competitive advantage in colonization and this may be why ST117 and ST80 have become dominant in their collection. We did not identify any homology to previously described *E. faecium* bacteriocins on the pELF backbone (63); however, pELF plasmids do encode multiple toxin-antitoxin systems that likely contribute to stabilizing the plasmid in the population (17,29,33). Vancomycin carriage in Tn*1546* is frequently co-located with toxin-antitoxin systems on circular plasmids (71), and examining the frequency of toxin-antitoxin systems on pELF backbones over time might give clues to the maintenance and dissemination of pELF plasmids in recent years.

The data completeness for all epidemiological variables ranged from 90 to 100 %, which is a strength of this study considering the longitudinal nature of this surveillance program. Unfortunately, data about antimicrobials administered in the last 30 days prior to isolate collection was not collected consistently and so was excluded from our analysis. Including antimicrobial use history in future analyses may explain the occurrence of some linezolid-resistant isolates.

We clustered all available pELF plasmids using MOB-suite and pling and found that in all major secondary clusters and subcommunities, there were pELF plasmids that did not contain AMR genes clustered with pELF plasmids that did. This further supports the idea that the pELF backbone is potentially a high-risk vector for AMR gene acquisition and horizontal spread (1,85).

We described the largest pELF plasmid to date at 330 kb, which is much larger than other reported pELF plasmids (average across NCBI: 114.8 kb). This type of co-integrate pELF plasmid was described previously in Hashimoto [et al 2025], and they gave three examples where the plasmids inserted into the pELF backbone were 19.3 kb, 39.4 kb, and 66.4 kb in size. We identified the insertion location of a circular ∼237 kb plasmid into the pELF backbone mediated by IS*1261E* insertion sequences, which indicates that larger pELF co-integrates are possible. IS*1216E* is known to insert through a targeted conservative mechanism (86), and the multiple copies of IS*1216E* found on most pELF plasmids provides opportunities for high-frequency co-integration and for shedding of extra cargo genes (19,30,33).

We demonstrated using short reads to screen for pELF plasmids, indicating that genomic surveillance of pELF plasmids is possible. However, there are still limitations with short-read data as the genomic location of AMR genes cannot be confirmed, and therefore long-read sequencing is required for resolving AMR gene carriage on pELF plasmids. More long-read sequencing needs to be done to further determine the contribution of linear plasmids to AMR gene dissemination in VRE both locally and globally.

Assembly and annotation of linear pELF plasmids remain challenging with our current bioinformatic tools, and this is likely why records of pELF plasmids have been infrequent until recently. The difficulty of identifying pELF-type plasmids was further reinforced by records in NCBI/PLSDB that were missing the hairpin or invertron sequences (**Table S3**), had extra complementary sequence past the hairpin, or were annotated as circular. Based on our experience, the choice of assembler had an impact on the final linear plasmid sequence; for example, Hybracter (48) and Unicycler (87) could assemble the majority of Pelf plasmid sequence, but they often did not capture the complete hairpin sequence. An example of this is illustrated in Almeida-Santos *et al*. (31); they used Unicycler for assembly, and their plasmids are missing hairpin sequences. Because of the assembler chosen, we are skeptical that they identified novel ends as their discussion suggests and unfortunately their sequencing read data was not publicly available for confirmation. In contrast, Flye (49) effectively captured the hairpin but often contained excess complementary sequence. Autocycler (47) supports linear contigs and could generate a complete pELF consensus sequence with some manual trimming and cleaning as described in the documentation and others have also shown this is an effective method to assemble pELF plasmids [Hall 2026]. This is our recommended assembly method if long read sets are sufficiently deep.

Altogether, this study furthers knowledge of pELF-type plasmids by demonstrating that linear pELF plasmids are relatively common across VRE blood isolates and by providing more examples of co- localized vancomycin and linezolid AMR genes on the same linear plasmid backbone. Co-localization of vancomycin and linezolid resistance genes on the same linear plasmid backbone has potential for rapid dissemination among *Enterococcus* isolates and merits monitoring in surveillance programs.

## Data Availability

All data produced in the present study are available upon reasonable request to the authors. WGS data is available on NCBI.

## ACKNOWLEDGEMENTS

We gratefully acknowledge Ken Fakharuddin for ONT sequencing, the Genomics Core Facility of the National Microbiology Laboratory, Public Health Agency of Canada for whole genome Illumina sequencing, the Bioinformatics Core Facility of the National Microbiology Laboratory, Public Health Agency of Canada, for computational infrastructure, and the Robotics Core Facility, National Microbiology Laboratory, Public Health Agency of Canada, for DNA extractions. We would like to acknowledge the members of the CNISP VRE working group: Jessica Bartoszko, Kelly Choi, John Conly, Jennifer Ellison, John M. Embil, George Golding, Susy S. Hota, Jennie Johnstone, Kevin Katz, Xena Li, Ben Mack, Melissa McCracken, Jennifer Parsonage, Stephanie W. Smith, Kathy N. Suh, Jen Tomlinson. We’d also like to thank the physicians, epidemiologists, infections control practitioners and laboratory staff at each participating hospital for their contributions to this study.

## REFERENCES

1. Almeida-Santos AC, Novais C, Peixe L, Freitas AR. Vancomycin-resistant *Enterococcus faecium*: A current perspective on resilience, adaptation, and the urgent need for novel strategies. Journal of Global Antimicrobial Resistance. 2025 Mar 1;41:233–52. doi:10.1016/j.jgar.2025.01.016

2. Ahmed MO, Baptiste KE. Vancomycin-Resistant Enterococci: A Review of Antimicrobial Resistance Mechanisms and Perspectives of Human and Animal Health. Microbial Drug Resistance. 2018 Jun 1;24(5):590–606. doi:10.1089/mdr.2017.0147

3. World Health Organization. WHO bacterial priority pathogens list, 2024: bacterial pathogens of public health importance, to guide research, development and strategies to prevent and control antimicrobial resistance [Internet]. Geneva, Switzerland; 2024 [cited 2026 Mar 30]. Available from: https://iris.who.int/items/dbc5f2d3-6d22-46d3-881c-b5e8c119d6c4

4. Canadian Nosocomial Infection Surveillance pubrogram. Healthcare-associated infections and antimicrobial resistance in Canadian acute care hospitals, 2019–2023. Can Commun Dis Rep. 2025;51(6–7):249–69. doi:10.14745/ccdr.v51i67a04 PubMed PMID: 40861922; PubMed Central PMCID: PMC12372953.

5. Bi R, Qin T, Fan W, Ma P, Gu B. The emerging problem of linezolid-resistant enterococci. Journal of Global Antimicrobial Resistance. 2018 Jun 1;13:11–9. doi:10.1016/j.jgar.2017.10.018

6. Pfaller MA, Cormican M, Flamm RK, Mendes RE, Jones RN. Temporal and Geographic Variation in Antimicrobial Susceptibility and Resistance Patterns of Enterococci: Results From the SENTRY Antimicrobial Surveillance Program, 1997–2016. Open Forum Infect Dis. 2019 Mar 15;6(Supplement_1):S54–62. doi:10.1093/ofid/ofy344

7. Kent AG, Spicer LM, Campbell D, Breaker E, McAllister GA, Ewing TO, et al. Sentinel Surveillance reveals phylogenetic diversity and detection of linear plasmids harboring *vanA* and *optrA* among enterococci collected in the United States. Howden BP, editor. Antimicrob Agents Chemother. 2024 Nov 6;68(11):e00591–24. doi:10.1128/aac.00591-24

8. Markwart R, Willrich N, Eckmanns T, Werner G, Ayobami O. Low Proportion of Linezolid and Daptomycin Resistance Among Bloodborne Vancomycin-Resistant Enterococcus faecium and Methicillin-Resistant Staphylococcus aureus Infections in Europe. Front Microbiol. 2021 May 31;12. doi:10.3389/fmicb.2021.664199

9. Piezzi V, Gasser M, Atkinson A, Kronenberg A, Vuichard-Gysin D, Harbarth S, et al. Increasing proportion of vancomycin-resistance among enterococcal bacteraemias in Switzerland: a 6-year nation-wide surveillance, 2013 to 2018. Euro Surveill. 2020 Sep 3;25(35):1900575. doi:10.2807/1560-7917.ES.2020.25.35.1900575 PubMed PMID: 32885778; PubMed Central PMCID: PMC7472687.

10. Hammerum AM, Karstensen KT, Roer L, Kaya H, Lindegaard M, Porsbo LJ, et al. Surveillance of vancomycin-resistant enterococci reveals shift in dominating clusters from vanA to vanB Enterococcus faecium clusters, Denmark, 2015 to 2022. Euro Surveill. 2024 Jun 6;29(23):2300633. doi:10.2807/1560-7917.ES.2024.29.23.2300633 PubMed PMID: 38847117; PubMed Central PMCID: PMC11158013.

11. Guo Y, Ding L, Yang Y, Han R, Yin D, Wu S, et al. Multicenter Antimicrobial Resistance Surveillance of Clinical Isolates from Major Hospitals — China, 2022. China CDC Wkly. 2023 Dec 29;5(52):1155–60. doi:10.46234/ccdcw2023.217 PubMed PMID: 38164466; PubMed Central PMCID: PMC10757731.

12. Beh JQ, Daniel DS, Judd LM, Wick RR, Kelley P, Cronin KM, et al. Genomics to understand the global landscape of linezolid resistance in Enterococcus faecium and Enterococcus faecalis. Microbial Genomics. 2025;11(6):001432. doi:10.1099/mgen.0.001432

13. Azzam A, Elkafas H, Khaled H, Ashraf A, Yousef M, Elkashef AA. Prevalence of Vancomycin-resistant enterococci (VRE) in Egypt (2010–2022): a systematic review and meta-analysis. J Egypt Public Health Assoc. 2023 Apr 11;98(1):8. doi:10.1186/s42506-023-00133-9

14. Wardal E, Kuch A, Gawryszewska I, Żabicka D, Hryniewicz W, Sadowy E. Diversity of plasmids and Tn1546-type transposons among VanA Enterococcus faecium in Poland. Eur J Clin Microbiol Infect Dis. 2017;36(2):313–28. doi:10.1007/s10096-016-2804-8 PubMed PMID: 27752789; PubMed Central PMCID: PMC5253160.

15. Partridge SR, Kwong SM, Firth N, Jensen SO. Mobile Genetic Elements Associated with Antimicrobial Resistance. Clin Microbiol Rev. 2018 Aug 1;31(4):e00088–17. doi:10.1128/CMR.00088-17 PubMed PMID: 30068738; PubMed Central PMCID: PMC6148190.

16. Freitas AR, Tedim AP, Francia MV, Jensen LB, Novais C, Peixe L, et al. Multilevel population genetic analysis of vanA and vanB Enterococcus faecium causing nosocomial outbreaks in 27 countries (1986–2012). J Antimicrob Chemother. 2016 Dec 1;71(12):3351–66. doi:10.1093/jac/dkw312

17. Johnson CN, Sheriff EK, Duerkop BA, Chatterjee A. Let Me Upgrade You: Impact of Mobile Genetic Elements on Enterococcal Adaptation and Evolution. Journal of Bacteriology. 2021 Oct 12;203(21):10.1128/jb.00177-21. doi:10.1128/jb.00177-21

18. Sadowy E. Linezolid resistance genes and genetic elements enhancing their dissemination in enterococci and streptococci. Plasmid. 2018 Sep 1;Antimicrobial Resistance and Mobile Genetic Elements99:89–98. doi:10.1016/j.plasmid.2018.09.011

19. Hall MB, Xue Y, Lee TSE, Herring E, Hume J, Wick RR, et al. Novel transposon Tn8026 acts as a global driver of transmissible linezolid resistance in Enterococcus via a linear plasmid [Internet]. medRxiv; 2026 [cited 2026 Mar 5]. p. 2026.03.04.26347163. Available from: https://www.medrxiv.org/content/10.64898/2026.03.04.26347163v1 doi:10.64898/2026.03.04.26347163

20. Cinthi M, Coccitto SN, Simoni S, Gherardi G, Palamara AT, Di Lodovico S, et al. The optrA, cfr(D) and vanA genes are co-located on linear plasmids in linezolid- and vancomycin-resistant enterococcal clinical isolates in Italy. Journal of Antimicrobial Chemotherapy. 2025 Mar 17;dkaf082.doi:10.1093/jac/dkaf082

21. Diaz L, Kiratisin P, Mendes RE, Panesso D, Singh KV, Arias CA. Transferable Plasmid-Mediated Resistance to Linezolid Due to cfr in a Human Clinical Isolate of Enterococcus faecalis. Antimicrobial Agents and Chemotherapy. 2012 Jun 14;56(7):3917–22. doi:10.1128/aac.00419-12

22. Li D, Li XY, Schwarz S, Yang M, Zhang SM, Hao W, et al. Tn6674 Is a Novel Enterococcal optrA-Carrying Multiresistance Transposon of the Tn554 Family. Antimicrobial Agents and Chemotherapy. 2019 Aug 23;63(9):10.1128/aac.00809-19. doi:10.1128/aac.00809-19

23. Hashimoto Y, Kita I, Suzuki M, Hirakawa H, Ohtaki H, Tomita H. First Report of the Local Spread of Vancomycin-Resistant Enterococci Ascribed to the Interspecies Transmission of a vanA Gene Cluster-Carrying Linear Plasmid. mSphere. 2020 Apr 8;5(2):10.1128/msphere.00102-20. doi:10.1128/msphere.00102-20

24. Hashimoto Y, Taniguchi M, Uesaka K, Nomura T, Hirakawa H, Tanimoto K, et al. Novel Multidrug-Resistant Enterococcal Mobile Linear Plasmid pELF1 Encoding vanA and vanM Gene Clusters From a Japanese Vancomycin-Resistant Enterococci Isolate. Front Microbiol. 2019 Nov 13;10. doi:10.3389/fmicb.2019.02568

25. Hashimoto Y, Suzuki M, Kobayashi S, Hirahara Y, Kurushima J, Hirakawa H, et al. Enterococcal Linear Plasmids Adapt to Enterococcus faecium and Spread within Multidrug-Resistant Clades. Antimicrobial Agents and Chemotherapy. 2023 Mar 28;67(4):e01619–22. doi:10.1128/aac.01619-22

26. Egan SA, Kavanagh NL, Shore AC, Mollerup S, Samaniego Castruita JA, O’Connell B, et al. Genomic analysis of 600 vancomycin-resistant Enterococcus faecium reveals a high prevalence of ST80 and spread of similar vanA regions via IS1216E and plasmid transfer in diverse genetic lineages in Ireland. J Antimicrob Chemother. 2022 Feb 1;77(2):320–30. doi:10.1093/jac/dkab393

27. Boumasmoud M, Dengler Haunreiter V, Schweizer TA, Meyer L, Chakrakodi B, Schreiber PW, et al. Genomic Surveillance of Vancomycin-Resistant Enterococcus faecium Reveals Spread of a Linear Plasmid Conferring a Nutrient Utilization Advantage. mBio. 2022 Mar 28;13(2):e03771–21. doi:10.1128/mbio.03771-21

28. Bakthavatchalam YD, Puraswani M, Livingston A, Priya M, Venkatesan D, Sharma D, et al. Novel linear plasmids carrying *van*A cluster drives the spread of vancomycin resistance in *Enterococcus faecium* in India. Journal of Global Antimicrobial Resistance. 2022 Jun 1;29:168–72. doi:10.1016/j.jgar.2022.03.013

29. McCracken M, Lerminiaux N, Adam HJ, Baxter M, Karlowsky JA, Golding GR, et al. Enterococcus faecium harbouring vanA, optrA and cfr(D) on a linear plasmid in Canada: a CANWARD surveillance case. J Antimicrob Chemother. 2026 Feb 1;81(2):dkaf478. doi:10.1093/jac/dkaf478

30. Sun L, Chen Y, Hua X, Chen Y, Hong J, Wu X, et al. Tandem amplification of the vanM gene cluster drives vancomycin resistance in vancomycin-variable enterococci. J Antimicrob Chemother. 2020 Feb 1;75(2):283–91. doi:10.1093/jac/dkz461

31. Almeida-Santos AC, Tedim AP, Duarte B, Silva LM, Teixeira J, Castro AP, et al. Unnoticed spread of linear VanA-plasmids in vancomycin-variable Enterococcus faecium strains across different regions: a diagnostics challenge. J Antimicrob Chemother. 2026 Jan 1;81(1):dkaf409. doi:10.1093/jac/dkaf409

32. Fujiya Y, Harada T, Sugawara Y, Akeda Y, Yasuda M, Masumi A, et al. Transmission dynamics of a linear vanA-plasmid during a nosocomial multiclonal outbreak of vancomycin-resistant enterococci in a non-endemic area, Japan. Sci Rep. 2021 Jul 20;11(1):14780. doi:10.1038/s41598-021-94213-5

33. Hashimoto Y, Dao DT, Kasuga I, Takemura T, Abe H, Hasebe F, et al. Ongoing independent evolution of linezolid and vancomycin-resistance pELF-type linear plasmids across the One Health spectrum. Antimicrobial Agents and Chemotherapy. 2025 Nov 18;69(12):e01168–25. doi:10.1128/aac.01168-25

34. Kurushima J, Ota N, Yoshii Y, Tomie N, Tomita H. A Novel Conjugation System in AMR-associated pELF-type Linear Plasmids of Enterococcus faecium [Internet]. bioRxiv; 2026 [cited 2026 Feb 5]. Available from: https://www.biorxiv.org/content/10.64898/2026.01.27.701956v1.full

35. Canadian Nosocomial Infection Surveillance Program. Surveillance Protocol for Vancomycin Resistant Enterococcus Bloodstream Infections [Internet]. Public Health Agency of Canada; 2026. Available from: https://ipac-canada.org/wp-content/uploads/2026/02/2026_CNISP_VRE_protocol_EN.pdf

36. Clinical and Laboratory Standards Institute. Performance Standards for Antimicrobial Susceptibility Testing: Informational Supplement M100 ED36:2026. Clinical and Laboratory Standards Institute, Wayne, PA, USA; 2026.

37. European Committee on Antimicrobial Susceptibility Testing. Breakpoint tables for interpretation of MICs and zone diameters v.16.0 [Internet]. 2026. Available from: https://www.eucast.org

38. Langmead B, Salzberg SL. Fast gapped-read alignment with Bowtie 2. Nat Methods. 2012 Apr;9(4):357–9. doi:10.1038/nmeth.1923

39. Danecek P, Bonfield JK, Liddle J, Marshall J, Ohan V, Pollard MO, et al. Twelve years of SAMtools and BCFtools. Gigascience. 2021 Feb 1;10(2):giab008. doi:10.1093/gigascience/giab008

40. Wick R. Porechop [Internet]. 2018 [cited 2023 Jan 3]. Available from: https://github.com/rrwick/Porechop

41. Steinig E, Coin L. Nanoq: ultra-fast quality control for nanopore reads. Journal of Open Source Software. 2022 Jan 8;7(69):2991. doi:10.21105/joss.02991

42. Krueger F, James F, Ewels P, Afyounian E, Weinstein M, Schuster-Boeckler B, et al. TrimGalore: v0.6.10 [Internet]. Zenodo; 2023 [cited 2023 Apr 5]. Available from: https://zenodo.org/record/7598955 doi:10.5281/zenodo.7598955

43. Andrews S. FastQC [Internet]. 2020 [cited 2023 Jan 3]. Available from: https://www.bioinformatics.babraham.ac.uk/projects/fastqc/

44. De Coster W, Rademakers R. NanoPack2: population-scale evaluation of long-read sequencing data. Bioinformatics. 2023 May 1;39(5):btad311. doi:10.1093/bioinformatics/btad311

45. Hall M. Rasusa: Randomly subsample sequencing reads to a specified coverage. JOSS. 2022 Jan 29;7(69):3941. doi:10.21105/joss.03941

46. Seemann T. Shovill [Perl] [Internet]. 2023 [cited 2023 May 29]. Available from: https://github.com/tseemann/shovill

47. Wick RR, Howden BP, Stinear TP. Autocycler: long-read consensus assembly for bacterial genomes. Bioinformatics. 2025 Aug 28;btaf474. doi:10.1093/bioinformatics/btaf474

48. Bouras G, Houtak G, Wick RR, Mallawaarachchi V, Roach MJ, Papudeshi B, et al. Hybracter: Enabling Scalable, Automated, Complete and Accurate Bacterial Genome Assemblies [Internet]. bioRxiv; 2023 [cited 2024 Jan 24]. p. 2023.12.12.571215. Available from: https://www.biorxiv.org/content/10.1101/2023.12.12.571215v1 doi:10.1101/2023.12.12.571215

49. Kolmogorov M, Yuan J, Lin Y, Pevzner PA. Assembly of long, error-prone reads using repeat graphs. Nat Biotechnol. 2019 May;37(5):5. doi:10.1038/s41587-019-0072-8

50. Wick RR, Schultz MB, Zobel J, Holt KE. Bandage: interactive visualization of de novo genome assemblies. Bioinformatics. 2015 Oct 15;31(20):3350–2. doi:10.1093/bioinformatics/btv383

51. Hall M. Resolving LRE linear plasmids [Internet]. 2026 Feb 13. doi:10.5281/zenodo.18626625

52. Bouras G, Grigson SR, Papudeshi B, Mallawaarachchi V, Roach MJ. Dnaapler: A tool to reorient circular microbial genomes. Journal of Open Source Software. 2024 Jan 11;9(93):5968. doi:10.21105/joss.05968

53. Oxford Nanopore Technologies. Medaka [Python] [Internet]. Oxford Nanopore Technologies; 2023 [cited 2023 Aug 23]. Available from: https://github.com/nanoporetech/medaka

54. Wick RR, Holt KE. Polypolish: Short-read polishing of long-read bacterial genome assemblies. PLOS Computational Biology. 2022 Jan 24;18(1):e1009802. doi:10.1371/journal.pcbi.1009802

55. Bouras G, Judd LM, Edwards RA, Vreugde S, Stinear TP, Wick RR. How low can you go? Short-read polishing of Oxford Nanopore bacterial genome assemblies [Internet]. bioRxiv; 2024 [cited 2024 Mar 15]. p. 2024.03.07.584013. Available from: https://www.biorxiv.org/content/10.1101/2024.03.07.584013v1 doi:10.1101/2024.03.07.584013

56. Bharat A, Petkau A, Avery BP, Chen JC, Folster JP, Carson CA, et al. Correlation between Phenotypic and In Silico Detection of Antimicrobial Resistance in Salmonella enterica in Canada Using Staramr. Microorganisms. 2022 Feb;10(2):2. doi:10.3390/microorganisms10020292

57. Bortolaia V, Kaas RS, Ruppe E, Roberts MC, Schwarz S, Cattoir V, et al. ResFinder 4.0 for predictions of phenotypes from genotypes. J Antimicrob Chemother. 2020 Dec 1;75(12):3491–500. doi:10.1093/jac/dkaa345

58. Seemann T. mlst [Internet]. 2023 [cited 2023 Jan 3]. Available from: https://github.com/tseemann/mlst

59. Hasman H, Clausen PTLC, Kaya H, Hansen F, Knudsen JD, Wang M, et al. LRE-Finder, a Web tool for detection of the 23S rRNA mutations and the optrA, cfr, cfr(B) and poxtA genes encoding linezolid resistance in enterococci from whole-genome sequences. J Antimicrob Chemother. 2019 Jun 1;74(6):1473–6. doi:10.1093/jac/dkz092

60. Petkau A, Mabon P, Sieffert C, Knox NC, Cabral J, Iskander M, et al. SNVPhyl: a single nucleotide variant phylogenomics pipeline for microbial genomic epidemiology. Microb Genom. 2017 Jun 8;3(6):e000116. doi:10.1099/mgen.0.000116 PubMed PMID: 29026651; PubMed Central PMCID: PMC5628696.

61. de Been M, Pinholt M, Top J, Bletz S, Mellmann A, van Schaik W, et al. Core Genome Multilocus Sequence Typing Scheme for High-Resolution Typing of Enterococcus faecium. J Clin Microbiol. 2015 Dec;53(12):3788–97. doi:10.1128/JCM.01946-15 PubMed PMID: 26400782; PubMed Central PMCID: PMC4652124.

62. Altschul SF, Gish W, Miller W, Myers EW, Lipman DJ. Basic local alignment search tool. J Mol Biol. 1990 Oct 5;215(3):403–10. doi:10.1016/S0022-2836(05)80360-2 PubMed PMID: 2231712.

63. Tedim AP, Almeida-Santos AC, Lanza VF, Novais C, Coque TM, Freitas AR, et al. Bacteriocin distribution patterns in Enterococcus faecium and Enterococcus lactis: bioinformatic analysis using a tailored genomics framework. Applied and Environmental Microbiology. 2024 Sep 16;90(10):e01376–24. doi:10.1128/aem.01376-24

64. Robertson J, Bessonov K, Schonfeld J, Nash JHE. Universal whole-sequence-based plasmid typing and its utility to prediction of host range and epidemiological surveillance. Microb Genom. 2020 Sep 24;6(10):mgen000435. doi:10.1099/mgen.0.000435 PubMed PMID: 32969786; PubMed Central PMCID: PMC7660255.

65. Robertson J, Nash JHE. MOB-suite: software tools for clustering, reconstruction and typing of plasmids from draft assemblies. Microb Genom. 2018 Jul 27;4(8):e000206. doi:10.1099/mgen.0.000206 PubMed PMID: 30052170; PubMed Central PMCID: PMC6159552.

66. Redondo-Salvo S, Bartomeus-Peñalver R, Vielva L, Tagg KA, Webb HE, Fernández-López R, et al. COPLA, a taxonomic classifier of plasmids. BMC Bioinformatics. 2021 Jul 31;22(1):390. doi:10.1186/s12859-021-04299-x

67. Tonkin-Hill G, MacAlasdair N, Ruis C, Weimann A, Horesh G, Lees JA, et al. Producing polished prokaryotic pangenomes with the Panaroo pipeline. Genome Biology. 2020 Jul 22;21(1):180. doi:10.1186/s13059-020-02090-4

68. Frolova D, Lima L, Roberts LW, Bohnenkämper L, Wittler R, Stoye J, et al. Applying rearrangement distances to enable plasmid epidemiology with pling. Microbial Genomics. 2024;10(10):001300. doi:10.1099/mgen.0.001300

69. Siguier P, Perochon J, Lestrade L, Mahillon J, Chandler M. ISfinder: the reference centre for bacterial insertion sequences. Nucleic Acids Res. 2006 Jan 1;34(Database issue):D32–36. doi:10.1093/nar/gkj014 PubMed PMID: 16381877; PubMed Central PMCID: PMC1347377.

70. Molano LAG, Hirsch P, Hannig M, Müller R, Keller A. The PLSDB 2025 update: enhanced annotations and improved functionality for comprehensive plasmid research. Nucleic Acids Research. 2025 Jan 6;53(D1):D189–96. doi:10.1093/nar/gkae1095

71. Arredondo-Alonso S, Top J, McNally A, Puranen S, Pesonen M, Pensar J, et al. Plasmids Shaped the Recent Emergence of the Major Nosocomial Pathogen Enterococcus faecium. mBio. 2020 Feb 11;11(1):10.1128/mbio.03284-19. doi:10.1128/mbio.03284-19

72. Kim D, Kang DY, Choi MH, Hong JS, Kim HS, Kim YR, et al. Fitness costs of Tn1546-type transposons harboring the vanA operon by plasmid type and structural diversity in Enterococcus faecium. Annals of Clinical Microbiology and Antimicrobials. 2024 Jul 8;23(1):62. doi:10.1186/s12941-024-00722-2

73. Raad II, Hanna HA, Hachem RY, Dvorak T, Arbuckle RB, Chaiban G, et al. Clinical-Use-Associated Decrease in Susceptibility of Vancomycin-Resistant Enterococcus faecium to Linezolid: a Comparison with Quinupristin-Dalfopristin. Antimicrobial Agents and Chemotherapy. 2004 Sep;48(9):3583–5. doi:10.1128/aac.48.9.3583-3585.2004

74. Mortelé O, van Kleef–van Koeveringe S, Vandamme S, Jansens H, Goossens H, Matheeussen V. Epidemiology and genetic diversity of linezolid-resistant *Enterococcus* clinical isolates in Belgium from 2013 to 2021. Journal of Global Antimicrobial Resistance. 2024 Sep 1;38:21–6. doi:10.1016/j.jgar.2024.04.010

75. Bender JK, Fleige C, Funk F, Moretó-Castellsagué C, Fischer MA, Werner G. Linezolid Resistance Genes and Mutations among Linezolid-Susceptible Enterococcus spp.—A Loose Cannon? Antibiotics. 2024 Jan 18;13(1). doi:10.3390/antibiotics13010101

76. Shoji R, Maeda M, Yamaguchi K, Takuma T, On R, Ugajin K, et al. Resistance mechanisms and tedizolid susceptibility in clinical isolates of linezolid-resistant bacteria in Japan. JAC Antimicrob Resist. 2025 Jun 1;7(3):dlaf097. doi:10.1093/jacamr/dlaf097

77. Deshpande LM, Castanheira M, Flamm RK, Mendes RE. Evolving oxazolidinone resistance mechanisms in a worldwide collection of enterococcal clinical isolates: results from the SENTRY Antimicrobial Surveillance Program. J Antimicrob Chemother. 2018 Sep 1;73(9):2314–22. doi:10.1093/jac/dky188

78. McCracken M, Wong A, Mitchell R, Gravel D, Conly J, Embil J, et al. Molecular epidemiology of vancomycin-resistant enterococcal bacteraemia: results from the Canadian Nosocomial Infection Surveillance Program, 1999-2009. Journal of Antimicrobial Chemotherapy. 2013 Jul 1;68(7):1505–9. doi:10.1093/jac/dkt054

79. AL Rubaye M, Janice J, Bjørnholt JV, Kacelnik O, Haldorsen BC, Nygaard RM, et al. The population structure of vancomycin-resistant and -susceptible Enterococcus faecium in a low-prevalence antimicrobial resistance setting is highly influenced by circulating global hospital-associated clones. Microbial Genomics. 2023;9(12):001160. doi:10.1099/mgen.0.001160

80. Rodríguez-Lucas C, Fernández J, Raya C, Bahamonde A, Quiroga A, Muñoz R, et al. Establishment and Persistence of Glycopeptide-Resistant Enterococcus faecium ST80 and ST117 Clones Within a Health Care Facility Located in a Low-Prevalence Geographical Region. Microbial Drug Resistance. 2022 Feb 1;28(2):217–21. doi:10.1089/mdr.2021.0171

81. Weber A, Maechler F, Schwab F, Gastmeier P, Kola A. Increase of vancomycin-resistant Enterococcus faecium strain type ST117 CT71 at Charité - Universitätsmedizin Berlin, 2008 to 2018. Antimicrob Resist Infect Control. 2020 Jul 16;9(1):109. doi:10.1186/s13756-020-00754-1

82. Centre for Disease Control. COVID-19: U.S. Impact on Antimicrobial Resistance, Special Report 2022 [Internet]. Atlanta, Georgia, United States: U.S. Department of Health and Human Services; 2022 Jun [cited 2026 Mar 27]. Available from: https://stacks.cdc.gov/view/cdc/117915 doi:10.15620/cdc:117915

83. Lagadinou M, Michailides C, Chatzigrigoriadis C, Erginousakis I, Avramidis P, Amerali M, et al. Impact of the COVID-19 pandemic on vancomycin-resistant Enterococcus bloodstream infections: a 6-year study in Western Greece. Front Microbiol. 2025 Oct 24;16. doi:10.3389/fmicb.2025.1656334

84. Garretto A, Dawid S, Woods R. Increasing prevalence of bacteriocin carriage in a 6-year hospital cohort of *E. faecium*. Federle MJ, editor. J Bacteriol. 2024 Dec 19;206(12):e00294–24. doi:10.1128/jb.00294-24

85. Lipworth S, Matlock W, Shaw L, Vihta KD, Rodger G, Chau K, et al. The plasmidome associated with Gram-negative bloodstream infections: A large-scale observational study using complete plasmid assemblies. Nat Commun. 2024 Feb 22;15(1):1. doi:10.1038/s41467-024-45761-7

86. Harmer CJ, Hall RM. IS26 Family Members IS257 and IS1216 Also Form Cointegrates by Copy-In and Targeted Conservative Routes. mSphere. 2020 Jan 8;5(1):10.1128/msphere.00811-19. doi:10.1128/msphere.00811-19

87. Wick RR, Judd LM, Gorrie CL, Holt KE. Unicycler: Resolving bacterial genome assemblies from short and long sequencing reads. PLOS Computational Biology. 2017 Jun 8;13(6):e1005595. doi:10.1371/journal.pcbi.1005595

