## Supplementary Figure 1 and 2 for "Linear plasmid prevalence and linezolid resistance gene carriage in vancomycin-resistant *Enterococcus* in Canada from 2009-2024"

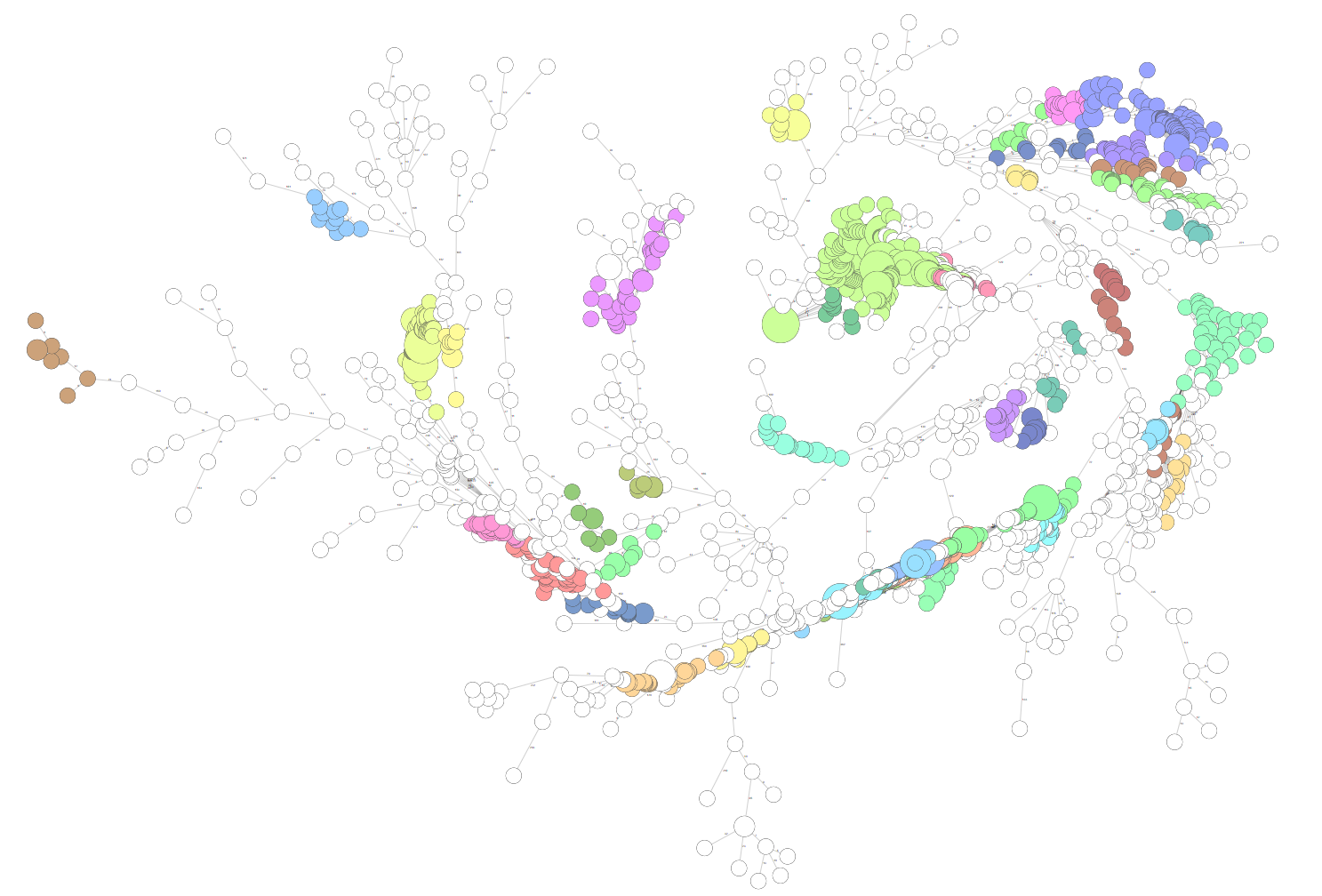

**Supplementary Figure 1**. Minimum spanning tree of core genome multi-locus sequence types (cgMLST) for n=2071 vancomycin-resistant *Enterococcus* (VRE) bloodstream infection isolates. The top 25 most common cgMLST types have coloured nodes in the plot, and other cgMLST type nodes are coloured white. Node size indicates number of isolates; bigger nodes indicate more isolates had identical cgMLST profiles.

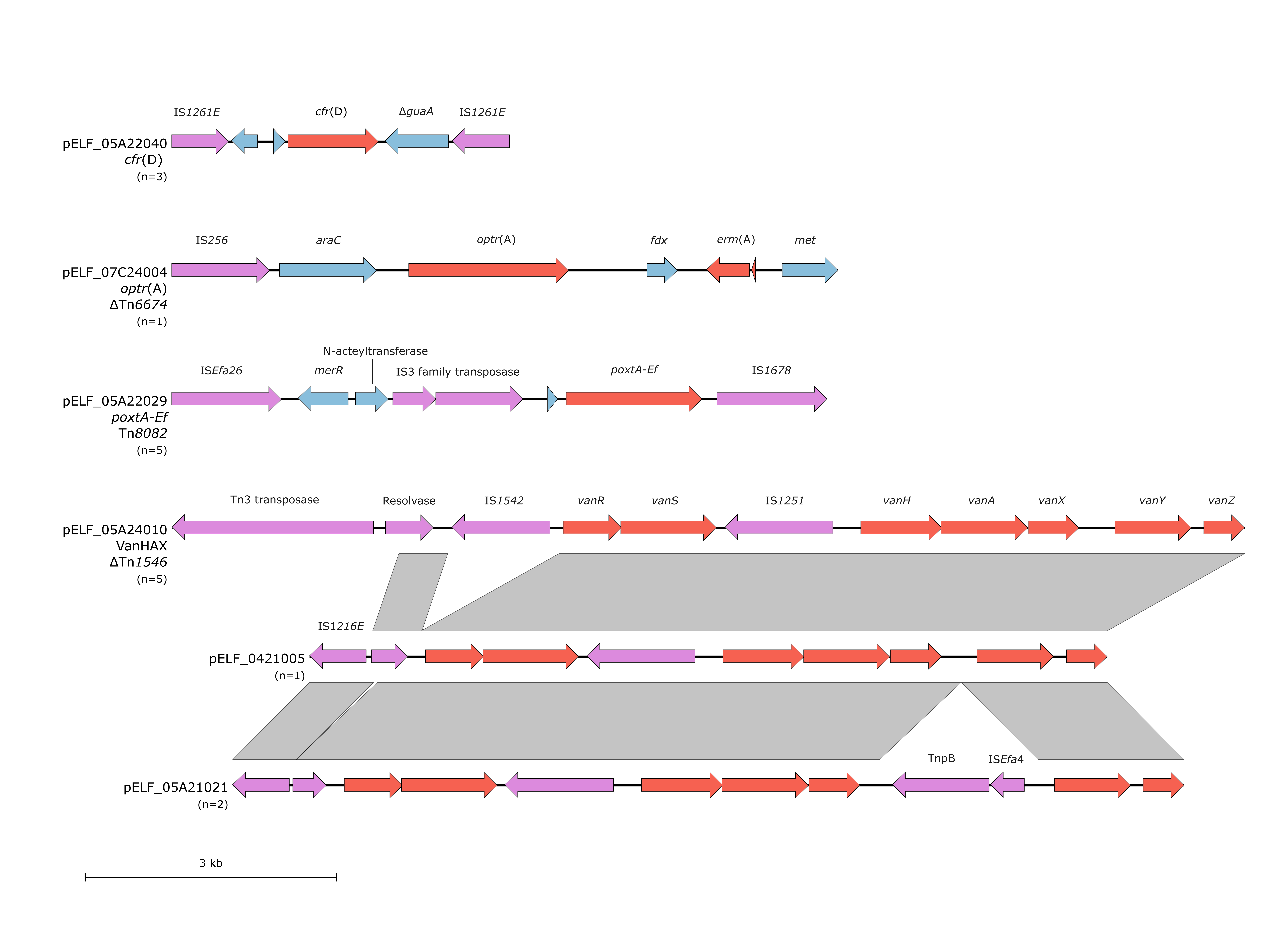

**Supplementary Figure 2.** Genetic context of *cfr*(D), *optr*(A), *poxtA*, and VanHAX on pELF linear plasmids. Genes are coloured by function: red are AMR genes, pink are mobile elements, and blue is other coding sequences. Grey blocks indicate 100% identity via blastn. A representative plasmid was chosen for the gene schematic and the total number of plasmids with this structure is represented by (n=x).
